# Automated Coronary Artery Disease Detection Using a CNN Model with Temporal Attention

**DOI:** 10.64898/2026.02.11.26346085

**Authors:** Keshav Balakrishna, Alessandro Hammond, Saanvi Cheruku, Adrito Das, Mehakpreet Saggu, Nidhi Abhay Thakur, Regina Urrea, Han Zhu

## Abstract

Coronary Artery Disease (CAD) is a leading cause of cardiovascular-related mortality and affects 20.5 million people in the United States and approximately 315 million people worldwide in 2022. The asymptomatic and progressive nature of CAD presents challenges for early diagnosis and timely intervention. Traditional diagnostic methods such angiography and stress tests are known to be resource-intensive and prone to human error. This calls for a need for automated and time-effective detection methods. In this paper, this paper introduces a novel approach to the diagnosis of CAD based on a Convolutional Neural Network (CNN) with a temporal attention mechanism. The model will be developed on an architecture that will automatically extract and emphasize critical features from sequential medical imaging data from coronary angiograms, allowing subtle signs of CAD to be easily spotted, which could not have been detected by convention. The temporal attention mechanism strengthens the ability of a model to focus on relevant temporal patterns, thus improving sensitivity and robustness in detecting CAD for various stages of the disease. Experimental validation on a large and diverse dataset demonstrates the efficacy of the proposed method, with significant improvements in both detection accuracy and processing time compared to traditional CNN architectures. The results of this study propose a scalable solution system for the diagnosis of CAD. This proposed system can be integrated into clinical workflows to assist healthcare professionals. Ultimately, this research contributes to the field of AI-driven healthcare solutions and has the potential to reduce the global burden of CAD through early automated detection.

## II. Introduction

In 2022, it was reported that more than 20.5 million U.S citizens and approximately 315 million people around the world have been reported to have CAD. [1]. Coronary artery disease occurs due to atherosclerosis, a blockage of the coronary arteries that supply blood to the heart muscle. Atherosclerosis can alter blood flow and lead to ischemia, which can result in severe complications, e.g., myocardial infarction, commonly known as heart attack, heart failure, and sudden cardiac death. Since CAD is insidious, symptoms do not usually appear until the disease has progressed to an advanced stage. [2].

Current CAD diagnosis methods rely on the use of one of three main imaging techniques: coronary angiography, computed tomography angiography (CTA), and magnetic resonance imaging (MRI). However, the problem with these approaches is that while they can be effective with advanced stages of CAD, these methods are invasive, expensive, and require specialized skillsets for accurate interpretation. For instance, coronary angiography involves the administration of contrast agents to coronary vessels which is not only an invasive procedure, but also carries risk of allergic reactions or arterial injuries. Noninvasive CTA and magnetic resonance imaging techniques only provide snapshots, and resolutions may be poor in both methods because small diseases, especially in their early stages, cannot be delineated. In addition Timely diagnoses of CAD are further complicated because reading such complex images is a subjective process which makes it prone to human error [3][4].

With growing demand for efficient and accurate CAD early detection methods, one promising solution is applying artificial intelligence (AI) and, specifically deep learning (DL) imaging technologies. Deep learning, especially the CNN model, has great potential in medical image analysis because it can automatically learn hierarchical features from raw images without any explicit feature extraction process, and it can discover many subtle patterns that are often undetectable by conventional methodologies. These benefits have made CNN the standard approach for a variety of image-based diagnosis-related tasks including the detection of lung cancer, diabetic retinopathy, and brain tumors [5][6].

However, CNN based CAD detection models still face some limitations. One of the key limitations of conventional CNNs is their inability to model temporal relationships in sequential medical imaging data. CAD often progresses slowly and requires the model to capture temporal dependencies to make accurate predictions. This temporal aspect is crucial, since early stage CAD might not show significant spatial features on an individual image; instead, the gradual deposition of plaque becomes evident once such features are analyzed over time. Therefore, it is essential for models that can integrate spatial and temporal features to provide an in-depth analysis of coronary artery disease progression.

In this regard, we propose a novel CNN architecture with a temporal attention mechanism. Temporal attention mechanisms are most commonly used in natural language processing and video analysis and focus on the most relevant parts of a sequence to capture long-range dependencies [7][8]. Incorporating temporal attention modules into CNN frameworks allow the most informative time-dependent features of coronary angiograms and provide better detection of early CAD, where temporal patterns are more indicative of disease progression than individual snapshots.

This paper proposes a deep learning model that implements a CNN model with the addition of a temporal attention mechanism for automatic analysis of sequential medical imaging data to detect CAD. Our model will identify and emphasize critical temporal features from coronary angiograms to enhance sensitivity and robustness of the CAD detection performance. We demonstrate extensive experiments on a large dataset of coronary angiograms to prove the efficacy of our approach by comparing the performance of our model with traditional CNN architectures and other state-of-the-art methods. Our results show significant improvement in detection accuracy.

## III. Literature Review

Cardiovascular diseases (CVDs) are responsible for approximately 31% of global deaths [1]. Early diagnosis and effective management are essential to minimize the prevalence and impact of these diseases. Recent advances in medical imaging with deep learning algorithms have revolutionized the detection and diagnosis of coronary artery disease (CAD) and other cardiovascular diseases.

### A. Deep Learning in Cardiovascular Imaging

Deep learning has shown promise to analyze medical images for cardiovascular disease. Al-Mallah et al. [4] noted that cardiovascular imaging plays an significant role in the diagnosis of CAD. Deep learning algorithms have significantly improved diagnostic accuracy and reduced the time taken for image interpretation. Zreik et al. [8] effectiveness of deep learning models in detecting cardiovascular disease from coronary angiography images. Their study employed a convolutional neural network (CNN) that outperformed the results of traditional methods by providing more accurate and faster diagnoses of coronary artery blockages.

In addition, advances in coronary CT angiography have also taken advantage of artificial intelligence in the detection of CAD. Zhang et al. [9] used deep learning to analyze coronary CT angiography scans and compare diagnostic performance to human experts. This was done in an effort to minimize reliance on invasive procedures.

### B. Global Health Burden and the Need for Enhanced Diagnostic Tools

Cardiovascular diseases require the development of improved diagnostic techniques. Murray et al. [2] presents global trends in mortality from registered deaths, and analyzes that CVD has significant disparities between regions. Murray’s findings explain that accessible diagnostic techniques are essential especially for low- and middle-income countries. Given these countries have limited healthcare infrastructure, deep learning’s implementation in cardiovascular diagnostics help address these challenges.

### C. Deep Learning for Coronary Angiography and Its Implications

Patel and Khera [3] discuss functionality of coronary angiography to detect blockages and assess severity of coronary artery disease. However, the interpretation of angiographic images requires highly trained specialists. Recent advances in deep learning algorithms, [6], have succeeded in the automation of the detection of diabetic retinopathy in retinal fundus photographs. These models could serve as a model for similar progress in cardiovascular imaging. By automating the analysis of coronary angiography images, deep learning models can help clinicians diagnose CAD more efficiently and accurately with less need for manual interpretation, thus allowing faster decision making.

## IV. Methodology

We present a CNN model made consisting of pre-trained Resnet18 architecture and the ability to detect coronary artery disease from a set of DICOM (Digital Imaging and Communications in Medicine) files.

**Fig. 1.**
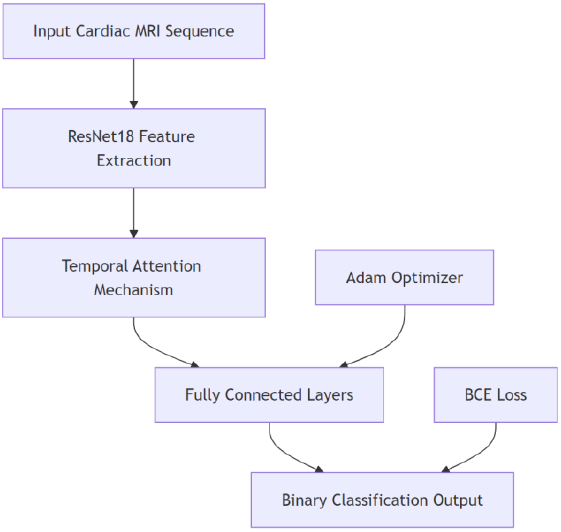
Resnet-18 Model Architecture

### A. Dataset

The training data for the model was obtained from a public Sunnybrook cardiac MRI dataset, which contained DICOM files of 10 different patients with a variety of pathologies: *healthy, heart failure, hypertrophy*, and *heart failure with infarction*. In total, 220 MRI images were used to train the input data.

## B. Algorithms

To enhance the adaptability of the program, the DICOM images were initially transformed into the Python Imaging Library (PIL) format. The pictures were then standardized to eliminate unnecessary information, or noise, that can impair model functionality. This pre-trained Resnet18 architecture employs 18 residual layers that leverage a direct correlation between input and output data, bypassing a number of modifications. As the layers learn from the input and output data, a non-linear mapping scheme is typically employed. To optimize the model, we implemented a Binary Cross Entropy (BCE) Loss Function as our criterion, and for 0 ≤ *y*_*i*_ ≤ 1:

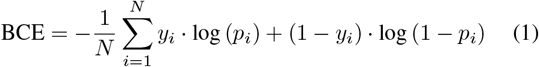

**Fig. 2.**
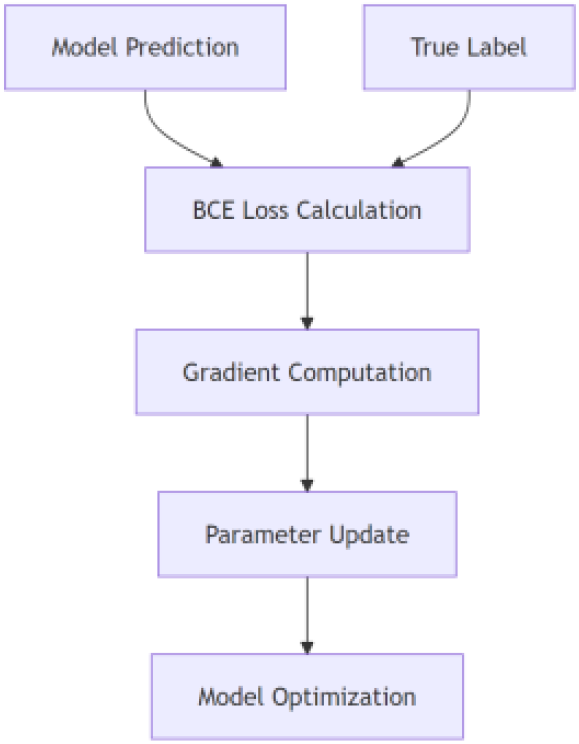
Diagram of Binary Cross Entropy Loss

*N* represents the number of samples, *y*_*i*_ represents the true label for the *i*-th sample, and *p*_*i*_ represents the predicted probability of the positive class for the *i*-th sample. For each sample, if *y*_*i*_ = 1, the loss is *−* log (*p*_*i*_), penalizing the model if *p*_*i*_ is small, and if *y*_*i*_ = 0, the loss is *−* log (1 *− p*_*i*_), penalizing the model if *p*_*i*_ is large. In this model, the loss was averaged over all *N* samples (MRI images) in the data set. BCE loss is an ideal choice for binary classification tasks, such as the one in this study, because it effectively penalizes incorrect predictions more harshly when the predicted probability is far from the true label, thus guiding the model learn better and achieve higher accuracy.

### C. Model Evaluation

The evaluation process used to assess the model consists of a loss calculation using BCE, a running loss aggregation, to accumulate the total loss, and a variety of performance metrics such as accuracy, precision, area under the curve (AUC), and recall.

We define the Running Loss as the discrete summation of all BCE loss of all batches (epochs) in the model’s training cycle. Mathematically, running loss can be defined as:

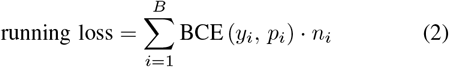

where *B* is the number of batches and *n*_*i*_ denotes the number of samples in the *i*-th batch. Afterward, the average loss is calculated and defined as:

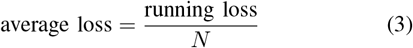

where *N* is the total number of samples in the dataset, providing the normalized measure of the performance of the model throughout the entire dataset, and thus offers a more comparable and stable metric across various dataset sizes and batch configurations.

### D. The Adam Optimizer

The Adam (Adaptive Moment Estimation) optimizer was implemented in the CAD detection model due to its efficacy in managing sparse gradients and irrelevant data, which are common in medical imaging tasks. Adam adjusts the learning rates for each individual parameter by using the estimates of the first and second gradient moments. This update rule for the Adam optimizer can be expressed as:

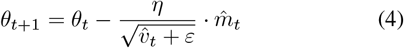

where *θ*_*t*_ is the parameter at time step *t, η* is the learning rate, 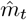 and 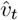 are the bias-corrected first and second moment estimates, respectively, and *ε* is a small constant to prevent division by zero.

The Adam optimizer is an effective combination of two optimization algorithms:

1. Momentum: It overcomes local minima and saddle points through an accumulation of previous weighted averages.
2. RMSprop: It adapts the learning rate for each parameter based on the averages of recent gradient magnitudes.

In the model, Adam was initialized to a learning rate of *η* = 0.001. This configuration allows the model to efficiently optimize its parameters, which was especially beneficial for the extraction of complex features from cardiac MRI images used for CAD detection.

### E. Temporal Attention Mechanism

*Def*. The temporal attention mechanism is a dynamic time selection mechanism that learns the long-time dependent characteristics of historical data and assigns higher-order weights to more relevant destinations over a given time period.

**Fig. 3.**
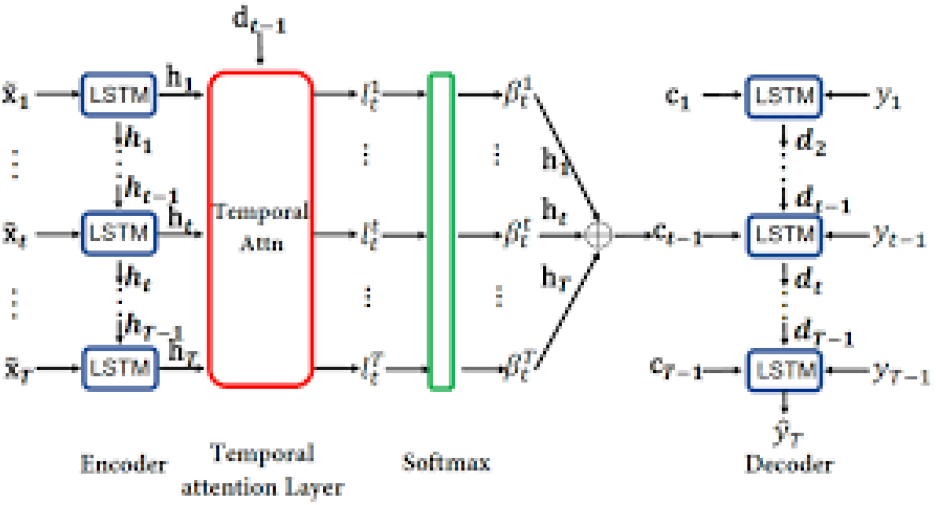
Example of Temporal Attention [12]

The temporal attention mechanism was a crucial component of the CNN model and improved its ability to focus on relevant temporal features across cardiac MRI image sequences. In general, the attention weights are computed using a linear transformation followed by a *softmax* function:

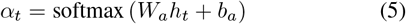

where *α*_*t*_ is the attention weight for time step *t, W*_*a*_ and *b*_*a*_ are learnable parameters, and *h*_*t*_ is the hidden state for time step *t*.

The softmax function has several applications in deep learning. Often used in the final layer of a neural network model for classification tasks, it converts raw output scores*−*also known as logits*−*into probabilities by exponentiating each output and normalizing each value by dividing the sum of all exponentials. From a linear algebra perspective, the function softmax takes a vector of *K* real numbers and transforms it into a probability vector of size *K* whose elements sum to 1.

At this point, we define the generation of the *context vector*. In attention, it is derived from the sum of attention weights, *α*_*t*_, multiplied by hidden states, *h*_*t*_:

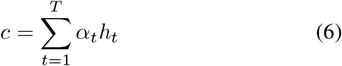

where *c* is the constant vector and *T* is the length of the sequence.

The temporal attention mechanism was integrated with the features extracted by the Resnet18 backbone. With the addition of Adam’s adaptive optimization with the selective focus of temporal attention, the Resnet18 architecture was suitable for the complex task of analyzing temporal patterns in cardiac MRI sequences for CAD detection.

## V. Results and Analysis

The CNN model achieved an accuracy score of 0.9787, or 97.87%, on the final training cycle, suggesting excellent performance in predicting CAD based on input data originating from the Sunnybrook dataset.

**Fig. 4.**
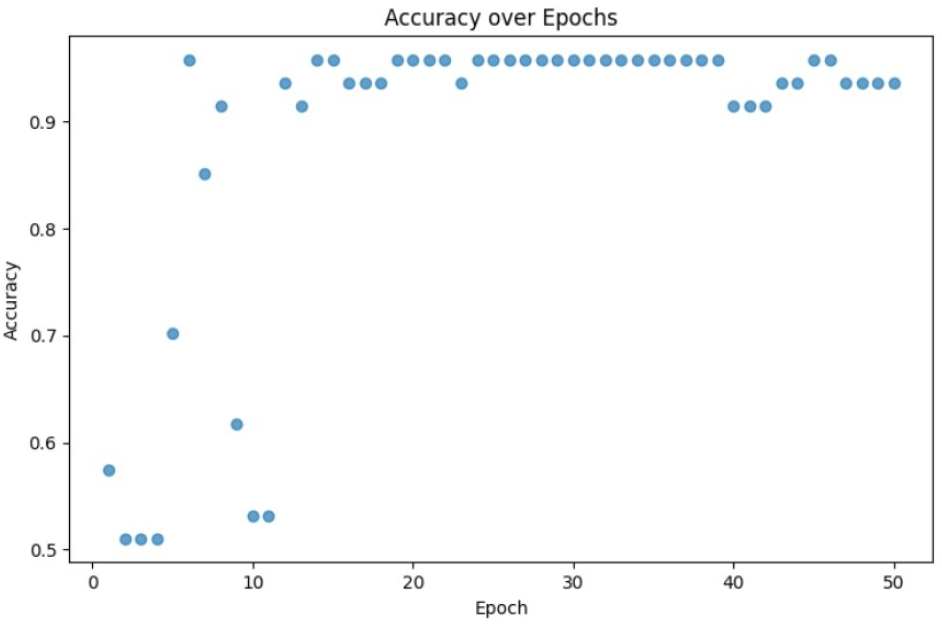
Graph of Accuracy Over the Training Cycle

The model had a precision score of [1.000 or] 100%, indicating its perfect accuracy in predicting the positive class, that is, it correctly identified all positive instances in the absence of false positives. Thus, the CNN model is ideal, as false positives in the diagnosis of this condition are highly undesirable.

**Fig. 5.**
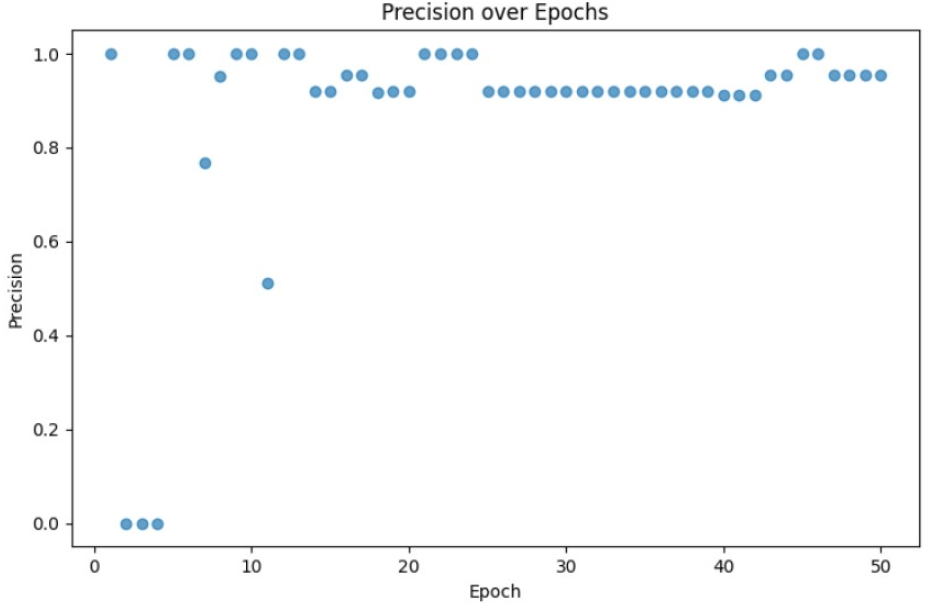
Graph of Precision Over the Training Cycle

The model had a recall of 0.9565, suggesting that 95.65% of the actual positive cases in the data were correctly identified. Thus, the most relevant instances of CAD were predicted correctly, which is significant as the model was very sensitive to true positive cases and underlines the effective nature of the Temporal Attention Mechanism.

**Fig. 6.**
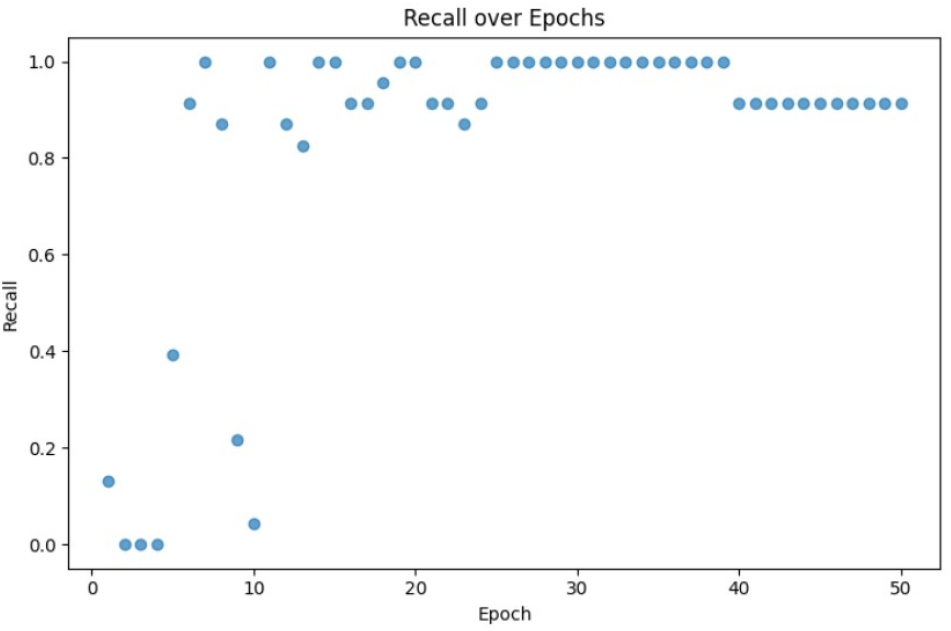
Graph of Recall Over the Training Cycle

The CNN model had an AUC (Area Under Curve) of 0.9982, or 99.82%. This suggested an exceptionally high level of performance, and that the model is able to distinguish positive and negative classes in CAD detection. This model performed a near-perfect classification with minimal room for error, close to the ideal score of 1.0000, which indicates a perfect classification.

**Fig. 7.**
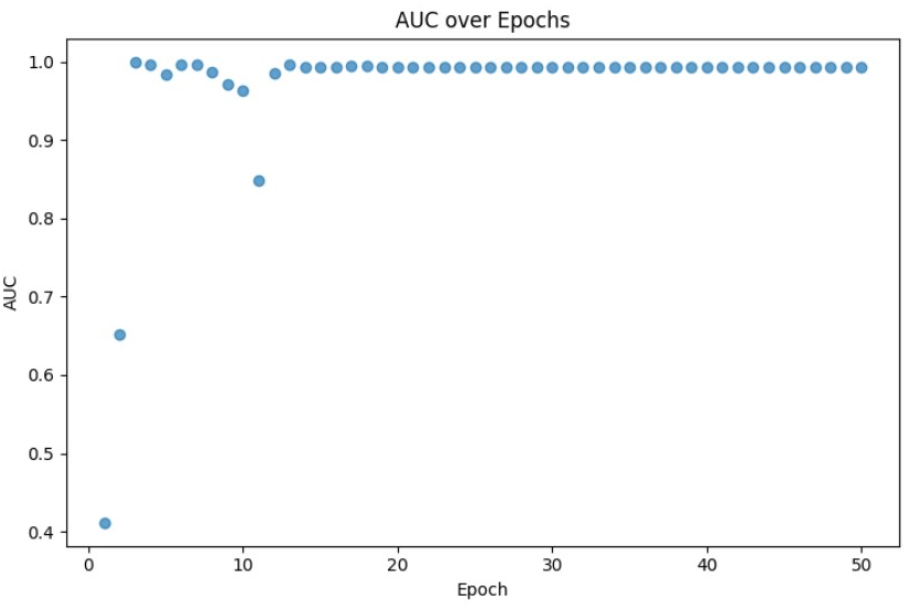
Graph of Area Under the Curve (AUC) Over the Training Cycle

Finally, the CNN model performed well in identifying legitimate cases of CAD in patients and not simply guessing the predictions. The line *y* = *x* on the Receiver Operating Characteristic (ROC) curve represents the probability of guessing randomly. After roughly 41 epochs in the training cycle, the data points are consistently above this line, close to the optimal value of 1.0.

**Fig. 8.**
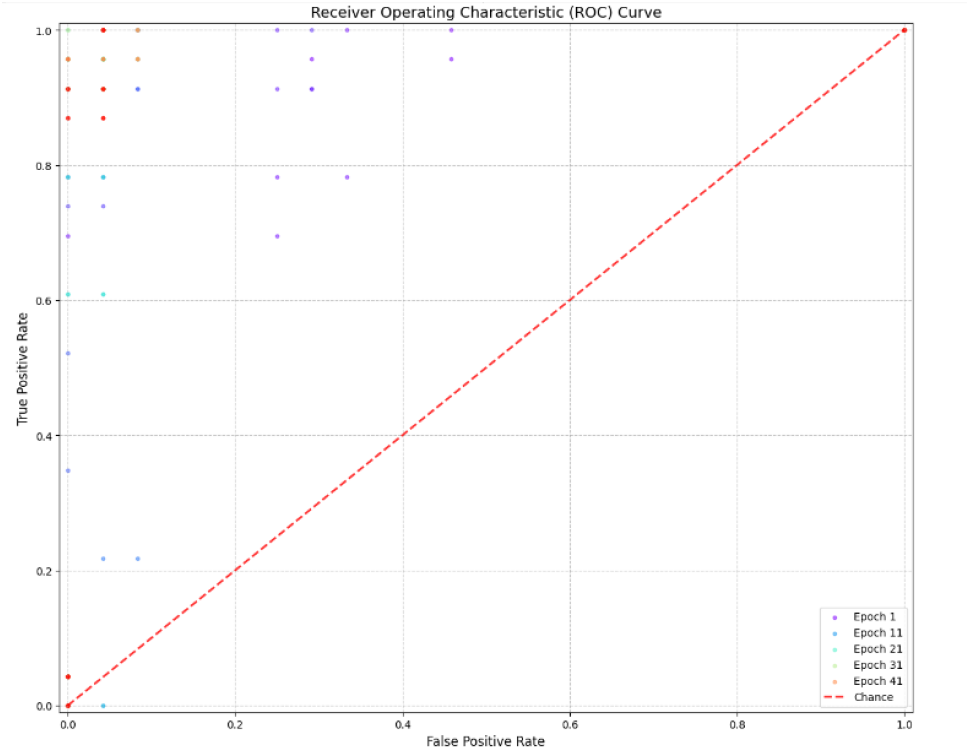
Graph of Receiver Operating Characteristic (ROC) Curve Over the Training Cycle

## VI. Conclusion

In this paper, we proposed an approach of a Convolutional Neural Network (CNN) architecture with a Temporal Analysis that can be used to predict Coronary Artery Disease (CAD) from a set of MRI images, provided by the public Sunnybrook dataset. Through the conversion of the images into DICOM files, data pre-processing, and augmentation using features such as normalization, and evaluation metrics such as binary cross-entropy (BCE) loss, the model delivered accurate and precise predictions of CAD and also demonstrated the potential of Machine Learning solutions for other medical conditions. Although the model illustrated positive results, additional features that incorporate more advanced data augmentation, e.g. elastic deformations and intensity adjustments (techniques often utilized for medical imagery), could have further improved the accuracy of the model. Furthermore, the application of model optimization techniques, such as model pruning and optimization, may have increased computational efficiency without sacrificing performance.

## Data Availability

All data produced in the present work are contained in the manuscript.

